# Diagnostic test accuracy of clinical scores in diagnosing Acute appendicitis in Right Iliac Fossa Pain

**DOI:** 10.1101/2024.03.30.24305104

**Authors:** Dev Desai, Rajan B. Patel, Jyot Patel

## Abstract

**Introduction:** For patients presenting to the Emergency room or OPD with acute severe right iliac fossa pain, the clinicians mostly prefer acute appendicitis as the first diagnosis, though many other differential diagnoses are there. Usually, in ER or OPD, clinical scores such as Alvarado Score, Ripasa scoring, and Tzanakis Scoring to diagnose Acute appendicitis. All these scores have different overlapping parameters and variable amounts of weightage. It is important to understand how sensitive and specific these scores are to determine the most accurate score to accurately diagnose acute appendicitis.

**Objectives:** Determine the Diagnostic accuracy of Alvarado Scoring, Ripasa Scoring, and Tzanakis Scoring compared to the Gold Standard Histopathology report.

**Methodology:** A Retrospective observational design was formulated where data of patients coming to the ER and OPD of the affiliated tertiary care hospital with right iliac fossa pain were enrolled. From the data, all clinical scores were calculated as per the standard guideline and the results of the scores were correlated with the gold standard Intraoperative Surgeon’s opinion and the patient’s histopathological report to determine the sensitivity, specificity, Youden Index and Accuracy of all the scores. Statistical software like EXCEL and SPSS along with Medcalc was used to produce the ROC Curve (Receiver Operating Characteristic Curve) & AUC of ROC and Fagan’s Nomogram.

**Results:** A total of 237 patients were enrolled who presented with right iliac fossa pain of which 156 were confirmed from the Histopathology report as patients with Acute Appendicitis. The Sensitivity was calculated for Alvarado (0.487), Ripasa (0.641), and Tzanakis(0.8269) whereas the Calculated Specificity was for Alvarado(0.8148), Ripasa(0.8395), Tzanakis(0.9699). The Younden Index was highest for Tzanakis score at 0.7968.

**Conclusion:** The Tzanakis Score is the most accurate Scoring method to diagnose Acute Appendicitis in patients presenting with RIF pain as it includes Ultrasonography. When USG is not available, the Ripasa score is the best scoring method while we do not recommend the use of Alvarado scoring due to poor sensitivity.

## Introduction

Acute appendicitis is one of the most common emergencies in the field of surgery. If not treated promptly may lead to perforation, abscess, and peritonitis eventually leading to Death of the patient. The definitive treatment of Acute appendicitis is by surgical removal i.e. appendicectomy for which we require prompt and accurate diagnosis. The diagnosis of appendicitis sometimes becomes challenging since the patients are usually in severe pain and due to guarding, patients don’t allow the doctor to perform necessary examinations. (1)

Acute appendicitis patients usually present with pain in the Right iliac fossa with vomiting. It is the most important screening feature and every doctor would be correct to put appendicitis as the first provisional diagnosis. To confirm the diagnosis, Ultrasonography results are widely considered when the inflamed appendix is visible and thus, the diagnosis becomes fairly simple. But when the appendix is not visible or when the diagnostic modality of USG is not available, it becomes far too crucial to diagnose the cause of Right iliac fossa pain. Here comes the role of clinical scores such as Alvarado, Ripasa, or Tzanakis which are widely employed by surgeons for the diagnosis of right iliac fossa pain even when USG is available. (2) (3) (4)

There can be multiple causes of Right iliac fossa pain other than appendicitis namely, Tiflitis, Ileocolitis, Renal calculi, or any other pathology. (5) It becomes important to diagnose the cause of the pain so the treatment modality can be confirmed. With proper analgesics and prompt pain relief, clinical investigations become easier, and hence, different Clinical scores become easier to employ. These clinical scores use different signs and symptoms to create a cut-off score above which it can be safely said that the patient is suffering from Acute appendicitis while scores lower than the cut-off prompt the surgeon to look for other causes of Right iliac fossa pain or use other signs or Diagnostic modality to completely rule out acute appendicitis. (6)

It is important to understand these clinical scores and their accuracy because, in third-world countries where most of the population live in far away remote areas and do not have access to advanced diagnostic modalities at their disposal, it is important to timely diagnose the patients correctly who present with right iliac fossa pain so that they can be treated promptly and complications, mortality, and morbidity can be decreased as much as possible. (7)

Alvarado, Ripasa, and Tzanakis scorings are in wide use today despite their efficacy and efficiency being questionable. There is an ongoing debate in the scientific world where some research papers support the use of these scores while other research papers disapprove of their role. (8) There are a few papers that support the use of these scores if they are being used as additionals to a definitive diagnostic modality. There are very few papers that compare the diagnostic accuracy of all these scores in the same set of patients and compare them with each other to find the best. That answer is the most important of all.

## Methodology

A retrospective cohort study design was adopted and patients who presented with right iliac fossa in the last 4 years at tertiary care hospitals were identified from records and enrolled. The records were accessed after permission from IRB as well as authorities. Those records were digitalized and fields to calculate clinical scores such as Alvarado, Ripasa, and Tzanakis were populated and total scores were calculated. Patients were included based on the availability of histopathology reports confirming their final diagnosis and all the data to populate the fields of clinical scores. Those patient records were excluded that did not have complete data. The cut-off value of Alvarado (at >4), Ripasa, (at >7.5), and Tzanakis (at >8) were determined and the data was analyzed. Sensitivity and specificity along with other diagnostic accuracy parameters were calculated from the status of their clinical scores and their histopathology report. Software like Excel 16 and SPSS20 along with REVMAN 5.0 and STATA14 were used to create the results.

## Results

A total of 237 patients were enrolled from the records who presented at the hospital with right iliac fossa pain. Out of the 237 patients, as visible in Table 1, 146 were males and 159 of them had confirmed Acute appendicitis.

**Table 1:** Patient Distribution.

| Table 1:- Patient Distribution |  |  |  |  |
| --- | --- | --- | --- | --- |
|  | Histopathology +ve | Histopathology -ve |  | Gender Distribution |
| Male | 97 | 49 | Male | 146 |
| Female | 59 | 32 | Female | 91 |
|  | Confirm Acute Appendicitis | Confirm No Appendicitis |  |  |
| Disease Distribution | 156 | 81 |  |  |

From the data collected from the records of these patients, fields of Clinical Scores such as Alvarado, Ripasa, and Tzanakis were populated and the scores were calculated. As per the determined Cut off values of each score, All patients were given a positive or a negative status and that was used to calculate the True Positives, True Negatives, False Positives, and False Negatives for each score to populate a 2^*^2 Contingency table. (Table 2:-Alvarado) (Table 3:-Ripasa) (Table 4:-Tzanakis)

**Table 2:** Alvarado Score.

| Table 2:- Alvarado Score |  |  |
| --- | --- | --- |
|  | Histopathology +ve | Histopathology -ve |
|  | Confirm Acute Appendicitis | Confirm No AA |
| Alvarado +ve (>4) | 76 | 15 |
| Alvarado -ve (<4) | 80 | 66 |

**Table 3:** Ripasa Score.

| Table 3:- Ripasa Score |  |  |
| --- | --- | --- |
|  | Histopathology +ve | Histopathology -ve |
|  | Confirm Acute Appendicitis | Confirm No AA |
| Ripasa +ve (>7.5) | 100 | 13 |
| Ripasa -ve (<7.5) | 56 | 68 |

**Table 4:**
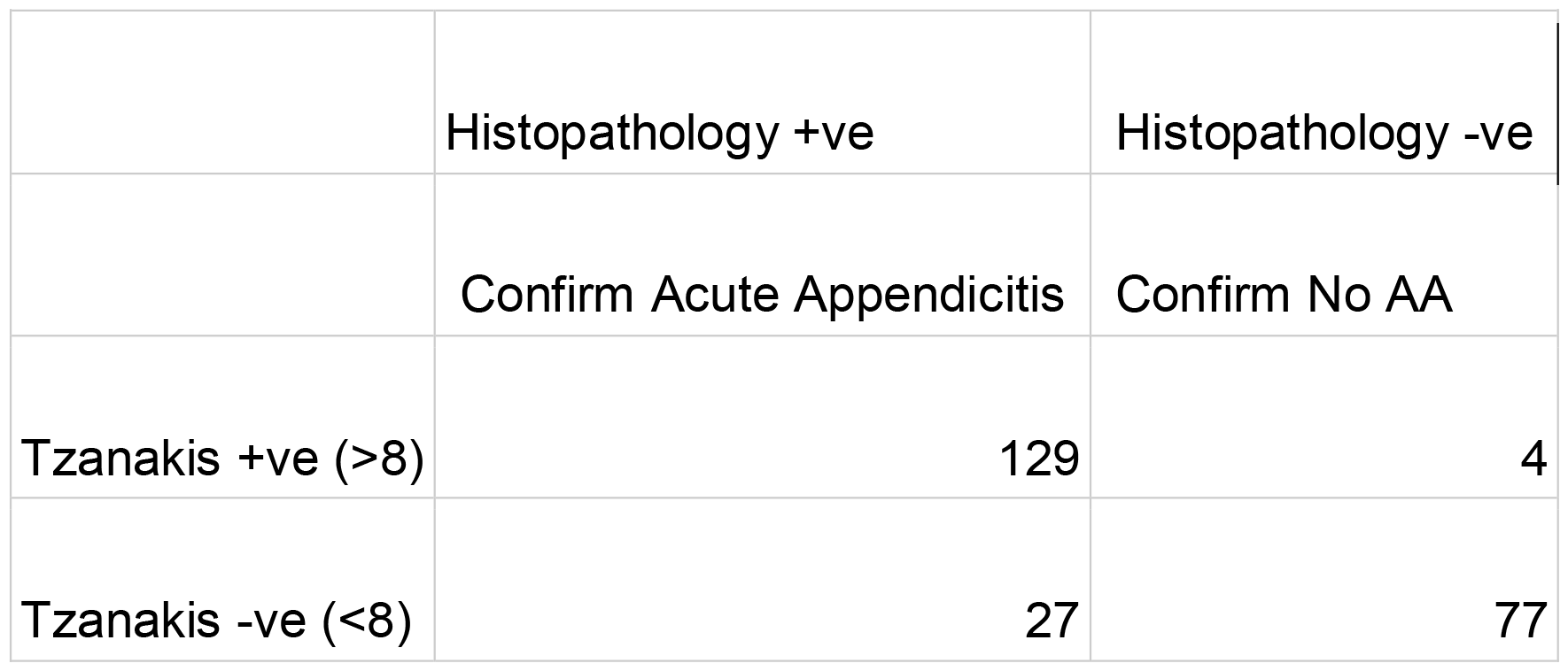
Tzanakis Score.

From these 2^*^2 Tables, Diagnostic test accuracy parameters were calculated for each score. (Table 5) along with this, ROC plots of each score were created to calculate the Area under the curve (AUC)

**Table 5:** Comparison of all Diagnostic parameters.

From these 2\*2 Tables, Diagnostic test accuracy parameters were calculated for each score. (Table 5)
| Table 5:- Comparison of all Diagnostic parameters |  |  |  |
| --- | --- | --- | --- |
|  | Alvarado | Ripasa | Tzanakis |
| Sensitivity | 0.487 | 0.641 | 0.827 |
| Specificity | 0.815 | 0.840 | 0.951 |
| PPV | 0.835 | 0.885 | 0.970 |
| NPV | 0.452 | 0.548 | 0.740 |
| LR +ve | 2.631 | 3.994 | 16.745 |
| LR -ve | 0.629 | 0.428 | 0.182 |
| Diagnostic Accuracy | 0.599 | 0.709 | 0.869 |
| DOR | 4.180 | 9.341 | 91.972 |

## Discussion

Acute appendicitis is a medical emergency that needs at least prompt pain relief and surgery otherwise the perforated bowel can lead to peritonitis, widespread infection, and inflammation and can even lead to death. In the area where Imaging modalities are not available, it is important to use the clinical scores and a surgeon needs to know the accuracy and precision of these scores to understand how much trust they can put while making a diagnosis and whether an urgent referral to tertiary care hospital is required or not.

The field has been studied extensively but it is still not widely known by surgeons and attending and it is always an issue for debate to decide which score to use in the time of need. This paper shows the problems with each score and why it should or should not be used.

It can be seen from Table 2, Table 5, and Figure 1 that Alvarado has very poor Sensitivity. At the Younden Index just sitting at 0.302, it can be safely said that Alvarado scoring pattern should not be implemented if any other scoring pattern is present. The Specificity and PPV make up for the lack of sensitivity with the values of 0.815 & 0.835, showing the useability of the test with a very low False Positive rate altogether. These findings of low Sensitivity were similar to Singla et al. 2016(9) and Korkut et al. 2020(6). The Specificity was as high as 1 in Singla et al. 2016 (9)and 0.91 in Nanjundaiah et al. 2014(10). The PPV of Alvarado has always been calculated to be above 0.75 in papers like Singla et al. 2016 (9)and Nanjundaiah et al. 2014(10). Alvarado shows poor NPV and Likelihood ratios bringing the AUC, Diagnostic Odds ratio, and Diagnostic accuracy down.

**Figure 1.**
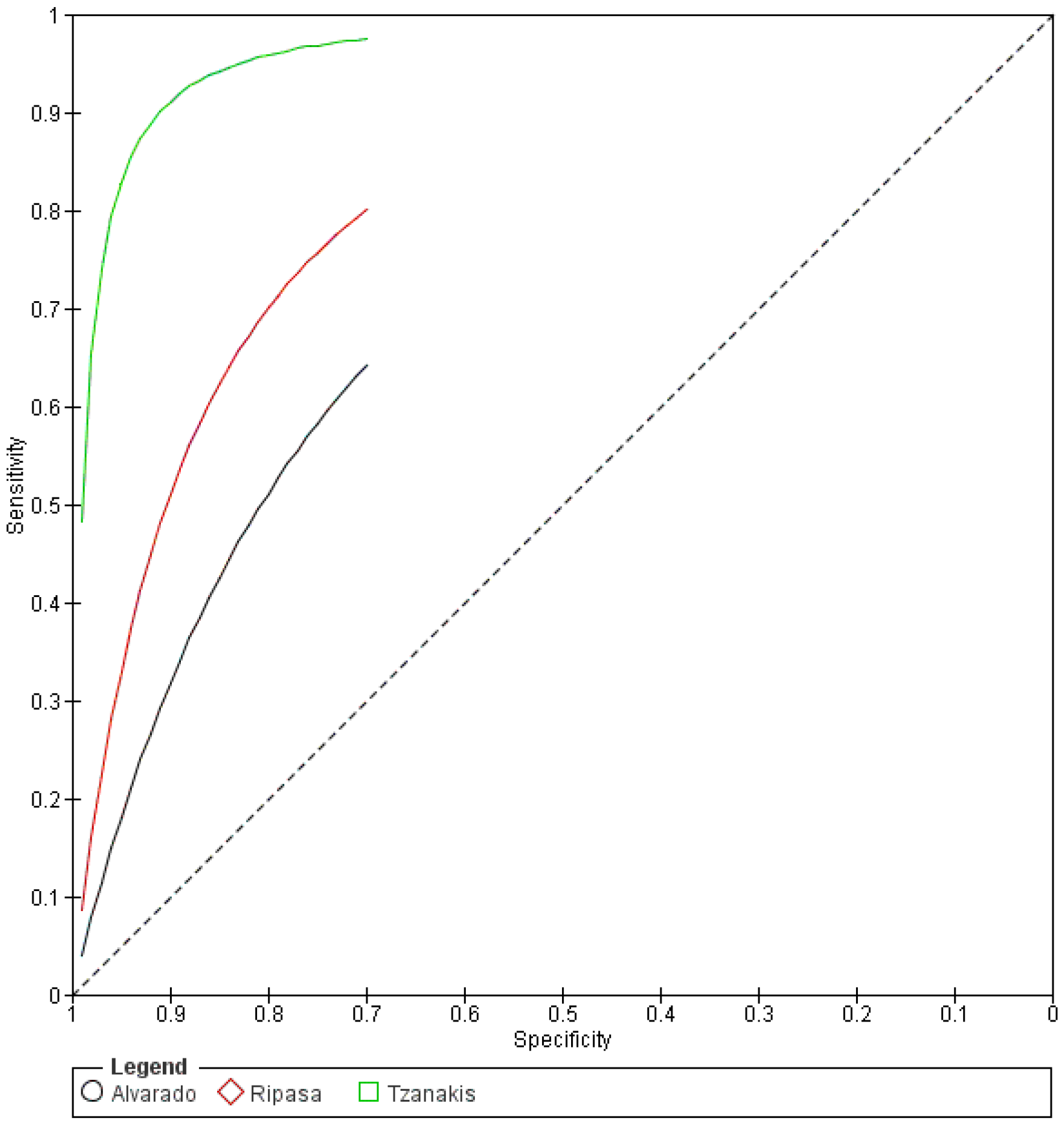
The area Under the Curve for Alvarado was 0.651, while for Ripasa, the AUC was 0.740; Tzanakis showed an AUC of 0.889. Along with that, the Younden Index for Alvarado was at 0.302, For Ripasa it was at 0.481 while for Tzanakis =, the Younden index was 0.778

In contrast to Alavarado, as visible in Table 3, Table 5, and Figure 1, Ripasa has a higher Sensitivity at 0.641 with similar Specificity and PPV similar to some previously done studies. Younden index sits at 0.481 which is better than Alvarado but still poor. Specificity was found to be at 0.84 similar to studies as Chong et al. 2011 (11) but several studies show a poor specificity below 0.5 such as Erdem et al. 2013(12). There are several discrepancies present in the scoring pattern of Ripasa as when it was formulated, a section for specific immigrants was included who at that time may have had a higher risk of getting acute appendicitis. This was known as NRIC criteria which is now not used anymore. Ripasa shows better NPV and Likelihood ratios bringing the AUC, Diagnostic Odds ratio, and Diagnostic accuracy higher than Alvarado.

Tzanakis is a scoring pattern that is not tested as much as Alvarado and Ripasa as the scoring pattern only has 4 questions. The question with the highest point asks about the presence of acute appendicitis changes on Ultrasonography and hence in the poor places where USG is not available at the surgeon or Medical officer’s disposal, the scoring pattern proves to be useless. However, because of this definitive Radiological imaging modality included in the Scoring, the sensitivity and specificity are very high with its Younden index sitting at 0.778 which is very good compared to Alvarado and Ripasa. This shows why Tzanakis has better NPV and Likelihood ratios along with AUC, Diagnostic Odds ratio, and Diagnostic accuracy higher than Alvarado and Ripasa.

## Conclusion

It can be seen from the results here that Tzanakis is the best scoring pattern available at the disposal of any surgeon or medical officer if USG is available. If USG is not available, Ripasa proves to be the best score while we recommend not using Alvarado scoring. Although considered one of the best scores, Alvarado score lacks diagnostic precision and cannot utilize the signs in the best manner possible.

## Data Availability

All data produced in the present study are available upon reasonable request to the authors

## Acknowledgement

The Authors would like to thank Smt. NHLMMC and SVPIMSR along with all the stakeholders for their support in this research paper. The authors would like to personally thank the following Surgery Postgraduate Resident doctors of the Department of Surgery at SVPIMSR hospital for the tremendous help in the completion of the project: Dr. Ankit, Dr. Drupad, Dr. Harshiv, Dr. Hardik, Dr. Krutarth, Dr. Muhammed, Dr. Chirag, Dr. Deep and Dr. Rutu.

